# DINASTY in NSCLC – a multicenter retrospective study on real-world data using federated analysis

**DOI:** 10.1101/2025.02.22.25321981

**Authors:** Å. Öjlert, P. Bhatnagar, L.E.L. Hendriks, F.R. Ogliari, F. Acker, E. Bolton, O. Bouissou, J. Brash, A.-L. Bynens, S. Cheeseman, H. Fenton, P. Galgane Banduge, A. Lobo Gomes, R. McDonald, P. Mahon, E. Ross, L. Sanchez Gomez, S. Theophanous, I. Zhovannik, Å. Helland

**Affiliations:** Deptartment of Oncology, Oslo University Hospital, Oslo, Norway; Non Surgical Oncology, Leeds Teaching Hospitals Trust, Leeds, United Kingdom; Pulmonary Diseases Department, GROW – Research Institute for Oncology and Reproduction, Maastricht University Medical Center, Maastricht, Netherlands; Department of Medical Oncology, IRCCS Ospedale San Raffaele, Milan, Italy; Department of Medicine II, Hematology and Oncology, Goethe University Frankfurt, University Hospital, Frankfurt Am Main, Germany; Research and Innovation Centre, Leeds Teaching Hospitals NHS, Leeds, United Kingdom; Department for Technology and E-health, Oslo University Hospital, Oslo, Norway; OMOP & PPG Solutions, IQVIA Ltd, London, United Kingdom; Clinical data science, Maastricht University Medical Center, Maastricht, Netherlands; Medical Oncology, Leeds Teaching Hospitals Trust, Leeds, United Kingdom; Oncology Evidence Network, IQVIA Ltd, London, United Kingdom; Department of Radiation Oncology (Maastro), Maastricht University Medical Center, Maastricht, Netherlands; Department of Radiation Oncology (Maastro), GROW Research Institute for Oncology and Reproduction, Maastricht, Netherlands; Real World Solutions, IQVIA Ltd, London, United Kingdom; DIGICORE, Bruxelles, Belgium; Federated Learning, Medical Data Works B.V., Maastricht, Netherlands; Research and Innovation, Leeds Teaching Hospitals NHS Trust, Leeds, United Kingdom; University of Oslo, Institute of Clinical medicine, Oslo, Norway

## Abstract

Real-world data is an important complement to randomized controlled trials and can be used to assess whether study results translate well to routine clinical practice and to groups of patients that are often excluded from clinical trials. The aim of this **di**sease **na**tural hi**st**ory and care qualit**y** assessment (DINASTY) study is to describe treatments, outcomes and care quality for patients with metastatic non-small cell lung cancer using real-world data. The study is a retrospective observational multicenter study conducted within DIGICORE, a non-profit European Economic Interest Grouping, formed to facilitate real-world evidence studies. The study will make use of methods developed within the network. Forty essential variables to describe patients with cancer, treatments and outcomes have been defined within DIGICORE and mapped to the Observational Medical Outcomes Partnership (OMOP) common data model (CDM). The study uses data that is drawn directly from the electronic patient health records at the patient’s local hospital and mapped to OMOP. Data are analyzed using a federated approach, meaning that patient-level data is analyzed locally, and only aggregated results are shared across centers and combined to present results for the full cohort. This method enables the delivery of multicenter studies and the presentation of results in a privacy-preserving way.

## Introduction

Randomized controlled trials are the gold standard for evaluating the effectiveness of cancer treatments, and they form the base for clinical practice guidelines. However, the results do not always translate well to a general patient population with cancer. Real-world evidence is therefore an important complement to clinical trials, both to validate the results of clinical trials in routine clinical practice and to assess clinical benefit in patients who would not have been eligible for the trial, e.g. due to comorbidities, age or performance status(1).

Lung cancer is a leading cause of cancer death globally(2). It is often diagnosed at an advanced stage, when treatment with curative intent is no longer possible. Non-small cell lung cancer (NSCLC) is the most prevalent subgroup of lung cancer(3). Several new treatments have been approved for NSCLC over the last few years, including immune checkpoint inhibitors and targeted therapies(4, 5). These treatments can give durable responses and long-term survival in a subset of patients, but it is still unclear whether the improvements found within a clinical study can be translated to a real-world patient population, to what degree patients with metastatic NSCLC are managed according to guidelines, and how to best use information on disease and patient characteristics to guide treatment decisions(6, 7).

DIGital Institute for Cancer Outcomes REsearch (DIGICORE; https://digicore-cancer.eu/) is a non-profit European Economic Interest Grouping with the aim to facilitate real-world studies. It consists of over 35 cancer centers and national cancer networks, in addition to two commercial partners. The group has reached a consensus on a Minimal Essential Description of Cancer (MEDOC) including 40 variables to describe the medical history, diagnosis, diagnostic work-up, treatment and outcomes of cancer patients(8). By gathering clinical information directly from the hospital’s systems, with possible manual or natural language enhancement of data on disease history, it is feasible to learn from every patient treated at that center. By assessing care quality objectives, legal implementation is simplified as such work counts as clinical audit, and so within the so called “primary use of data” (to enhance care delivery). We describe this combination as a <u>DI</u>sease <u>NA</u>tural hi<u>ST</u>ory and care qualitY assessment or DINASTY study. Federated analyses equivalent to centralized data analyses, can then be applied to gain insights from patient data collected at multiple cancer centers in a privacy-preserving way, without clinical patient-level data ever leaving the participating hospital. The DINASTY in NSCLC study is the first complex real-world study performed within the DIGICORE network and using the methods developed within the network. The aim of the study is to assess the natural history, treatment, outcomes, and guideline compliance in patients with metastatic NSCLC.

## Protocol

### Objectives

The primary objective is to describe the clinical and disease characteristics closest to the index date (date of diagnosis of metastatic NSCLC), re-biopsy rates, and anti-cancer therapies received for NSCLC prior to the index date in patients with metastatic NSCLC. Secondary objectives include to describe the 1^st^ and 2^nd^ lines of treatment (LoT) prescribed for metastatic NSCLC, and overall survival (OS) and time to next treatment by line of treatment. These analyses will be performed for all patients with metastatic NSCLC and in subgroups with metastases to the brain, liver, adrenal gland, bone, lung, other single sites, and multiple sites of metastases.

Additional secondary objectives include to describe duration of treatment and dose intensity according to body mass index, age and gender for patients receiving immunotherapy as their 1^st^ line of treatment, and to benchmark quality of care in the participating centers based on ESMO recommendations for diagnostic work-up and systemic treatment of metastatic NSCLC(4, 5). The primary and secondary objectives are listed in Table 1.

**Table 1.**
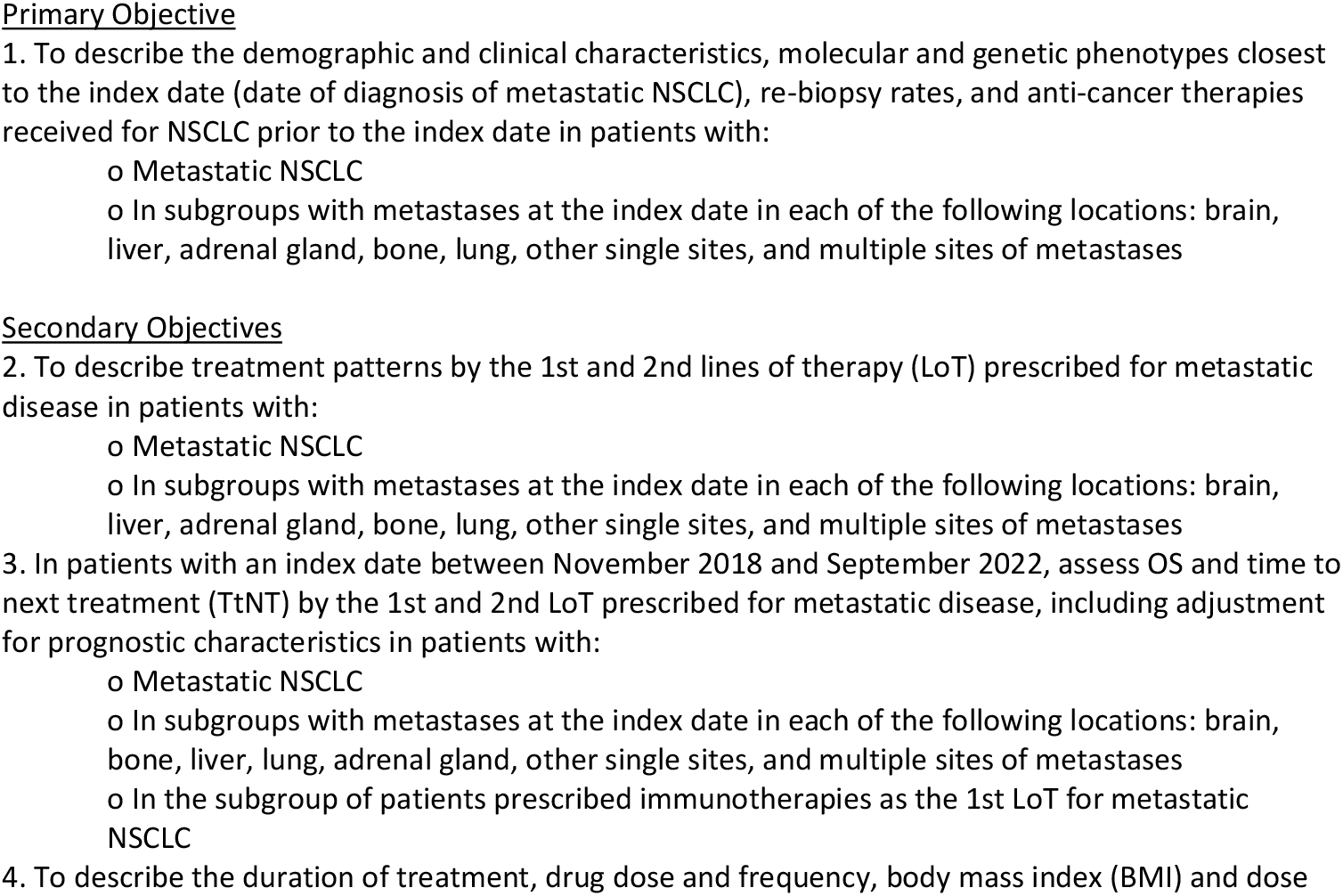
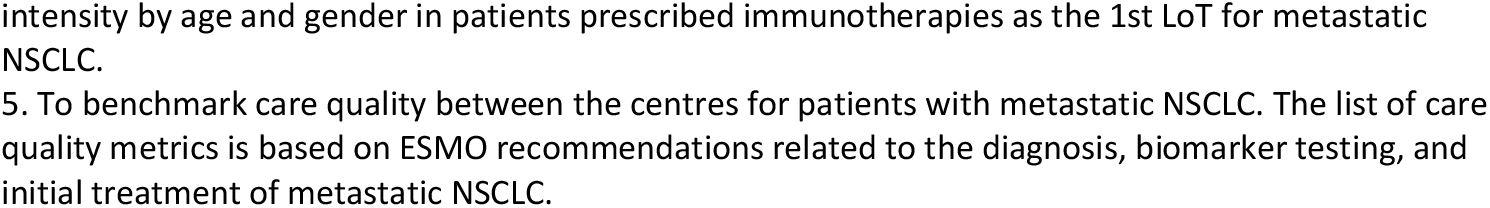

### Study design

This is a retrospective observational multi-center study describing the diagnostic work-up, treatment and outcomes for patients with metastatic NSCLC. Both patients with metastatic disease at diagnosis and patients who relapse after treatment for limited stage disease are eligible, as long as they have an index date between 1^st^ November 2018 and 30^th^ November 2023. The index date is defined as the date of diagnosis of metastatic NSCLC. Patients who recorded to opt out of data use for research purposes and patients for whom data necessary to assess the primary and secondary objectives is missing are excluded from the study. However, patients with sufficient information to assess some of the objectives can still be included in the analyses for these objectives.

OS and time to next treatment (TtNT) will only be analyzed in patients with an index date between 1^st^ November 2018 and 30^th^ September 2022, to allow for a minimum of 14 months follow up (Figure 1). Quality of care will both be assessed in the full cohort and in patients with an index date after 17th of January 2023, when the current ESMO guidelines were published.

**Figure 1.**
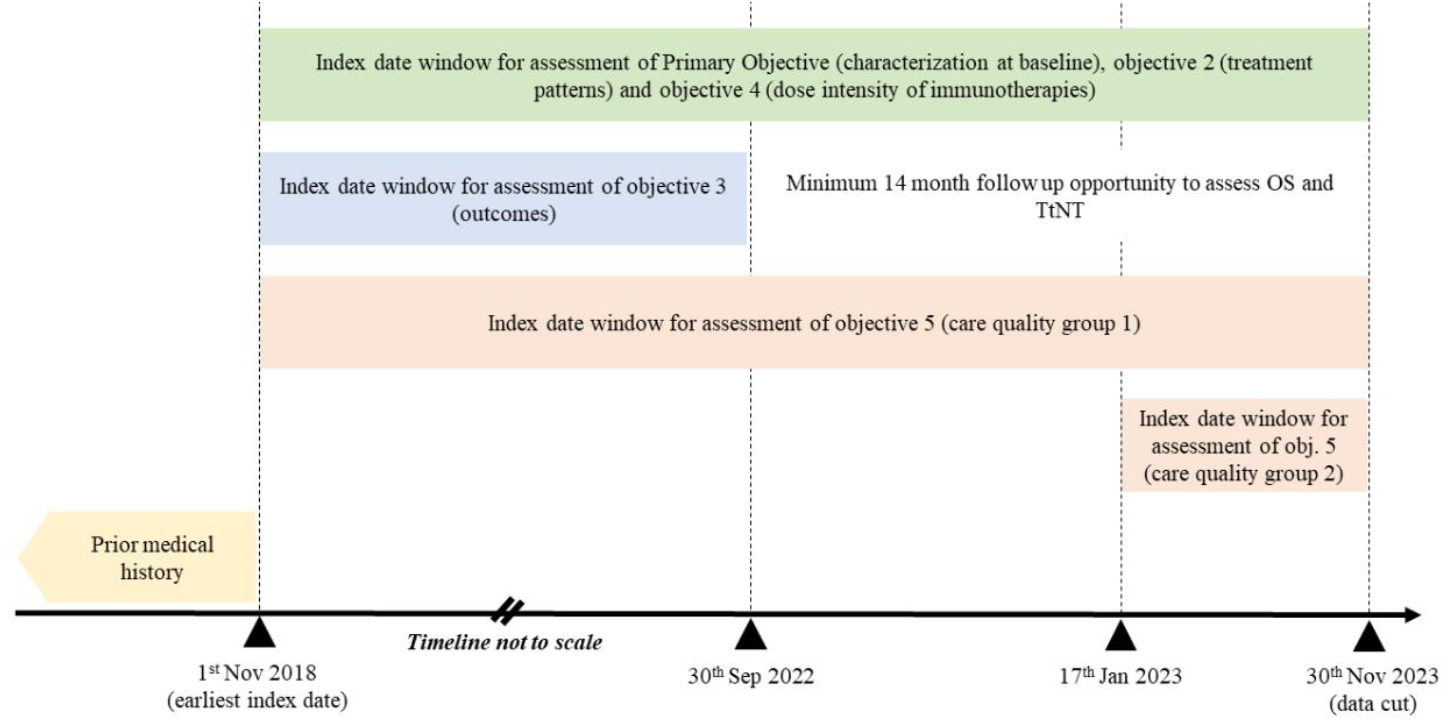
Schematic diagram of time windows

### Data sources

Five centers in the Digital Oncology Network for Europe (DigiONE) will participate in the study. These are located in Germany, Italy, the Netherlands, Norway, and the United Kingdom. The MEDOC variables that are to be used in the analyses will be collected from the electronic medical records and mapped to the Observational Medical Outcomes Partnership (OMOP) common data model (CDM) (https://www.ohdsi.org/data-standardization/). The date of death will be collected from the regional or national death registry, if possible. All data concepts that are required for the analysis are listed in Table 2. Additional centers may join the DIGICORE network and participate in this study. Requirements for the participation of additional centers include: data concepts listed in Table 2 accessible in OMOP CDM format; the ability to get the contractual agreements in place and set up the technical infrastructure to conduct federated analysis; and the ability to meet study timelines.

**Table 2.**
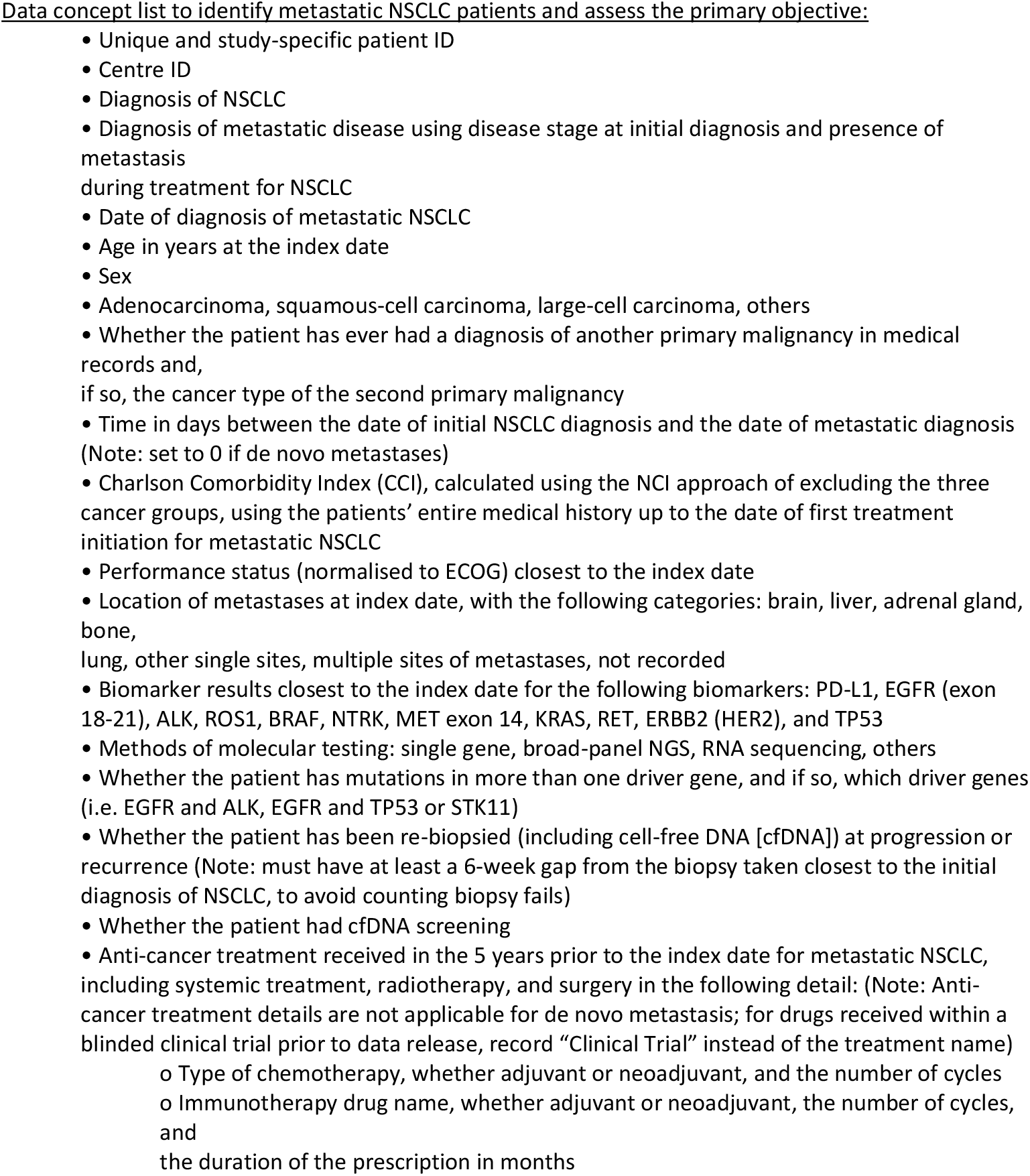
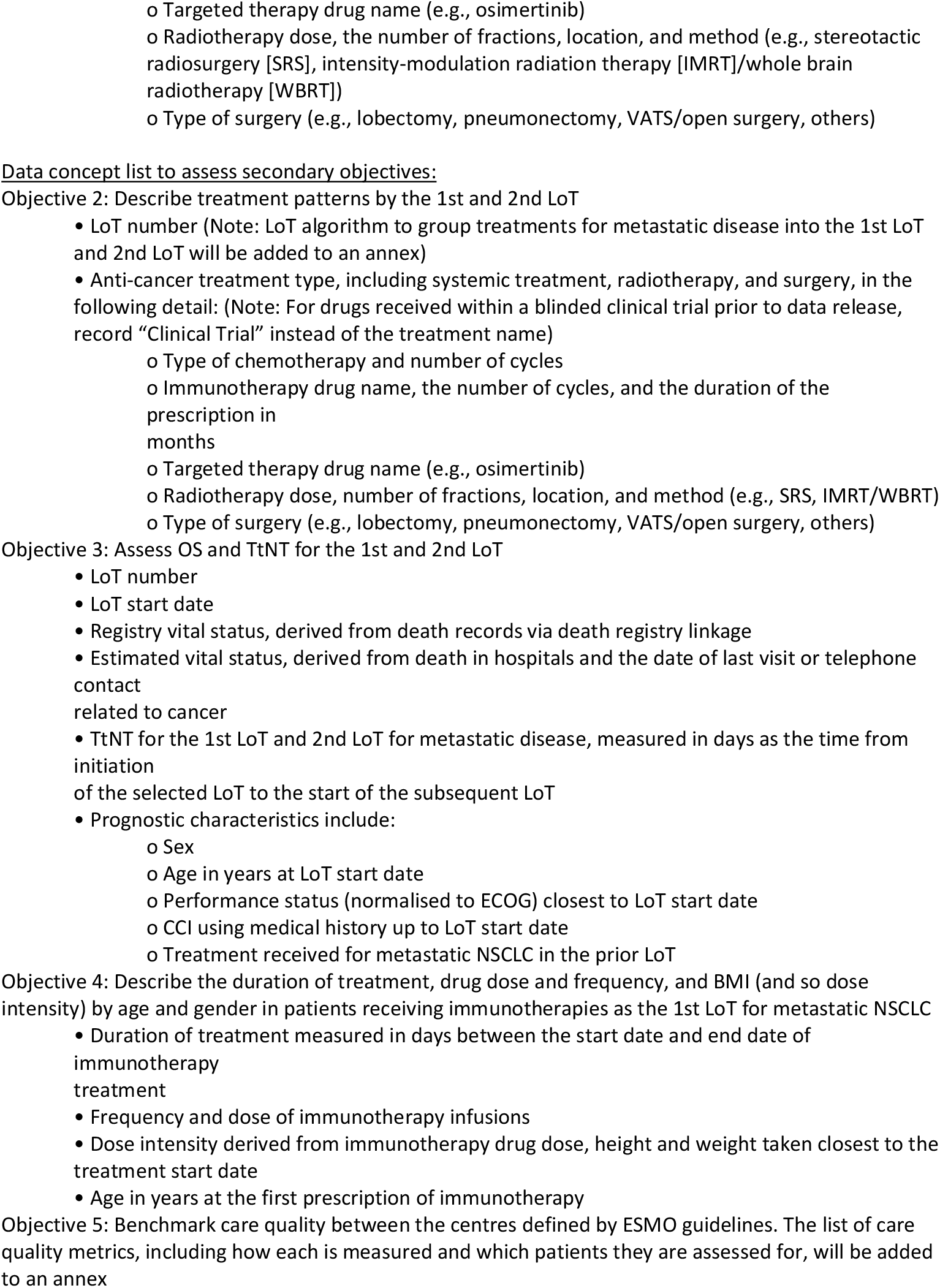
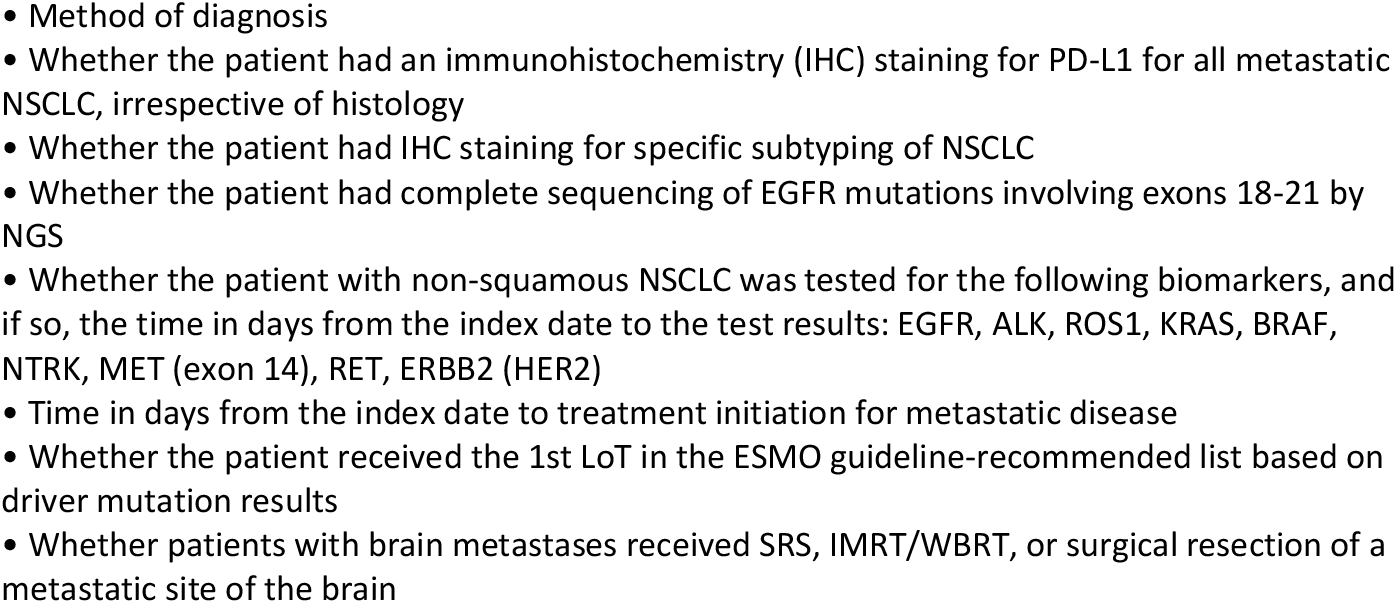

### Statistical analysis

Full details of the analysis approach and the table shells are documented in a statistical analysis plan (SAP).

The analyses in the study will be performed using a federated methodology. We also strive for robust privacy preservation by applying methods such as differential privacy. The combined methods lead to a privacy-preserving approach in which no patient-level data leaves the participating center. In short, scripts for performing the statistical analyses are developed centrally and shared with all participating centers. The analyses are carried out locally by running these scripts on patient data in the OMOP format. The results are then shared and used to merge and report results for the full cohort. The data flow in the federated network is illustrated in Figure 2. Each centers dataset is a node in the network and each node has a connection to a central server. The researcher using the central server cannot view the patient-level data at the nodes and each of the nodes cannot view the data at another node. Vantage 6 v2.1 is used to manage the network(9). As an additional measure to protect patient-level data, no patient count below five patients at each center will be reported; instead the entry ‘<5’ will be used. Secondary suppression will be employed so that the ‘<5’ cell cannot be calculated from other results.

**Figure 2.**
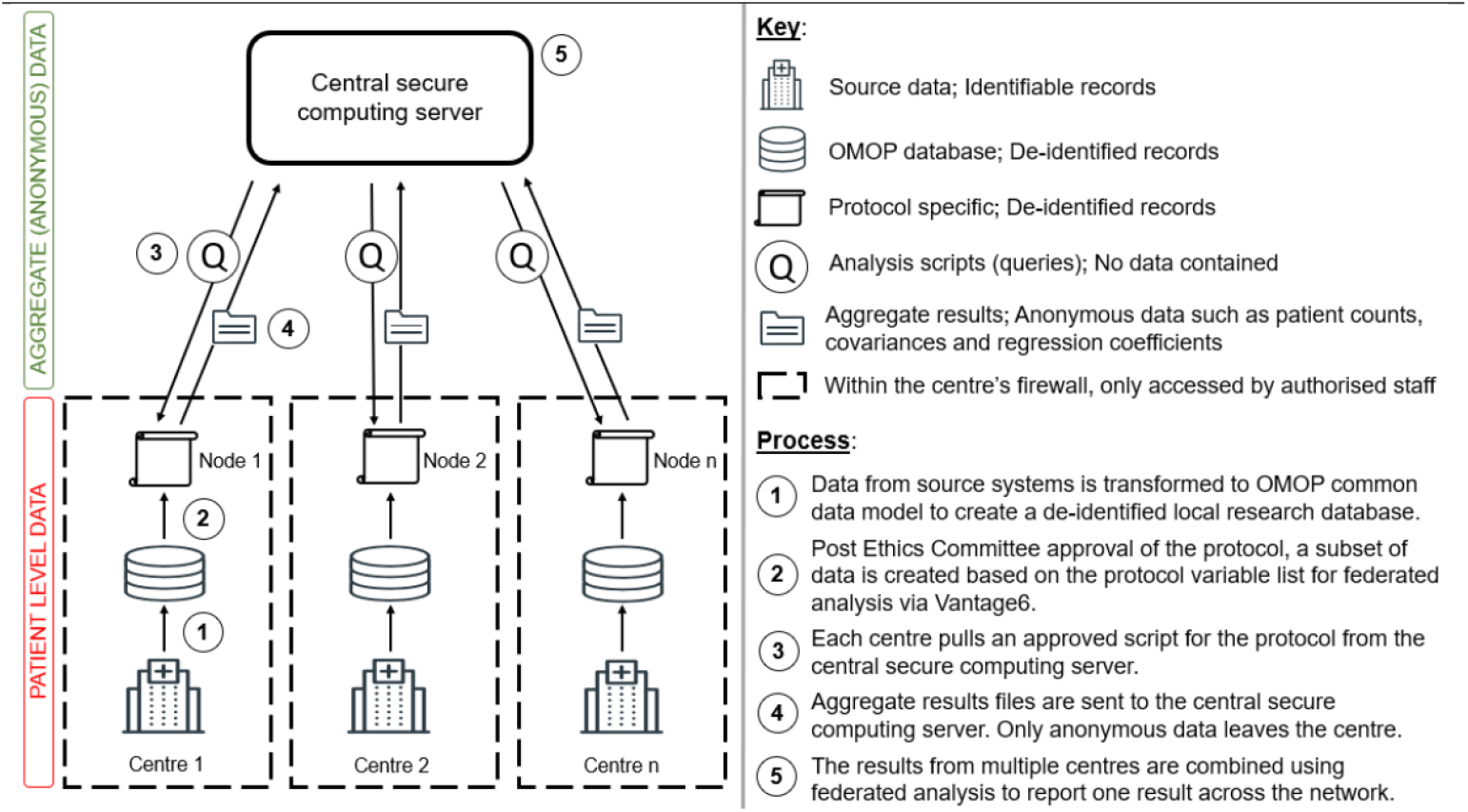
Data flow in a federated network

For continuous variables, the number of observations, mean, standard deviation (SD), median, first and third quartile, interquartile ranges [IQR]), minimum, and maximum will be provided. For categorical variables, the proportion of the total number of patients will be expressed in percentage and frequency in each category. The time-to-event endpoints OS and TtNT will be estimated using the Kaplan-Meier (KM) method and adjusted for the following variables, if available: sex, age in years at LoT start date, performance status closest to LoT start date, Charlson Comorbidity Index (CCI) using medical history up to LoT start date and treatment received for NSCLC in the prior LoT. The results of the analyses will be depicted graphically by KM curves and summarized by 25th, 50th, and 75^th^ percentiles with associated 95% confidence intervals (CIs) as well as the number and percentage of events, and number and percentage of censored observations.

A line of treatment algorithm will be utilized to describe the 1^st^ and 2^nd^ line of treatment received for metastatic NSCLC. ESMO guideline recommendations for the diagnosis and initial treatment of metastatic NSCLC will be used to assess care quality. These metrics will be reported for the whole cohort and per center. The center name and location will be blinded when these results are presented.

### Ethical considerations

This study is non-interventional; therefore, there is no change to the clinical standard of care as a result of this protocol. The analysis is based on the secondary use of existing data, and a specific informed consent module for patients is not required in this study. This protocol has been submitted to the relevant ethics committees per center for approval prior to the study’s commencement, unless there was an existing approval that covers this study’s design. This study will be conducted in accordance with the applicable laws and regulations of the region or country/countries where the study is being conducted.

## Data Availability

All data produced in the present study are available upon reasonable request to the authors and after ethical approval at the relevant institutions.

## Acknowledgements

The study is part of the DIGICORE research network, and only DIGICORE members can take part in the study.

